# Oxygen pulse kinetics and ventilatory inefficiency as markers of cardiovascular limitation on exercise in patients with mild pre-capillary pulmonary hypertension and exertional dyspnoea

**DOI:** 10.64898/2026.03.10.26347216

**Authors:** Athanasios Charalampopoulos, Sirpi Malar Selvaraju, Ian Smith, Ellis Cerrone, Rahul Mohanraj, Robin Condliffe, Charlie A. Elliot, Abdul G. Hameed, Judith A. Hurdman, Alexander M.K. Rothman, Andrew J. Swift, David G. Kiely, A. A. Roger Thompson

## Abstract

**Introduction:** Cardiopulmonary exercise testing (CPET) quantifies exercise limitation and helps differentiate cardiovascular dysfunction from deconditioning in patients with exertional dyspnoea. In mild pulmonary arterial hypertension (PAH) and chronic thromboembolic pulmonary hypertension (CTEPH), traditional CPET oxygen delivery parameters may not adequately distinguish cardiac limitation. We evaluated whether oxygen pulse (O₂ pulse) kinetics and the ratio of ventilation–carbon dioxide slope to peak oxygen uptake (VEVCO₂/peakVO₂) improve identification of cardiovascular limitation and prognostication.

**Methods:** We retrospectively analysed 289 consecutive patients referred for CPET. Patients were categorised into pre-capillary PH, no PH, or “unclassified” PH based on haemodynamics. O₂ pulse slopes were calculated across exercise phases, and qualitative curve patterns were classified. VEVCO₂/peakVO₂ was derived from standard CPET parameters. Logistic regression assessed predictors of cardiac dysfunction (peak O₂ pulse <65% predicted). Survival was evaluated using Kaplan–Meier and Cox regression analyses.

**Results:** Pre-capillary PH patients demonstrated more impaired aerobic capacity and ventilatory efficiency than those without PH. Abnormal O₂ pulse patterns (early plateauing or down-sloping) were associated with shallower slopes, lower peak O₂ pulse, and greater chronotropic index. A work-phase O₂ pulse slope < 0.40 identified impaired oxygen delivery but was not independently predictive in multivariable analysis. VEVCO₂/peakVO₂ independently predicted cardiac dysfunction (OR 3.9 [2.6-6.2], p < 0.001) and showed strong discrimination (AUC 0.83). VEVCO₂/peakVO₂ ≥ 2.7 independently predicted mortality (HR 13.6, 95% CI 3.8–48.5, p<0.001) outperforming peak O₂ pulse and VE/VCO₂ slope.

**Conclusion:** O_2_ pulse kinetics, particularly a work-phase slope < 0.40 and plateauing or decreasing trajectories, are associated with cardiac dysfunction in patients with pre-capillary PH. VEVCO₂/peakVO₂ appears to be a marker of cardiovascular limitation and mortality and may aid differentiation between cardiac dysfunction and deconditioning in this population when conventional CPET parameters are inconclusive.

## Introduction

Cardiopulmonary diseases are characterised by reduced exercise capacity and exertional dyspnoea. Cardiopulmonary exercise testing (CPET) can quantify aerobic exercise capacity, distinguish between respiratory and cardiovascular causes of dyspnoea, and assist risk stratification and treatment escalation (1–4). Oxygen delivery to the peripheral muscles during exercise can be impaired by cardiac failure, anaemia and reduced oxygen extraction pathologies including deconditioning and myopathies (5). The classic parameters of oxygen delivery on CPET are: oxygen consumption at the anaerobic threshold (AT), oxygen uptake (VO_2_) per workload (VO_2_/work), chronotropic index as a measure of heart rate response to exercise, and oxygen pulse (O_2_ pulse) (6). In pulmonary arterial hypertension (PAH) and in chronic thromboembolic pulmonary hypertension (CTEPH) remodelling of the small-sized pulmonary arteries and stenoses/occlusions due to chronic pulmonary emboli respectively can increase pulmonary vascular resistance (PVR) leading to right ventricular (RV) dysfunction and failure over time (7, 8). In patients with PH, deconditioning is not uncommon (9–11). In mild PH, both RV dysfunction and deconditioning may cause similar mild changes in oxygen delivery parameters (O_2_ pulse, AT, VO_2_/work) on exercise making the differentiation between the two challenging (12, 13).

O_2_ pulse represents the amount of oxygen extracted per heartbeat and is calculated as the ratio of VO_2_ to heart rate (HR) (14). The ratio VO_2_/HR is the product of stroke volume (SV) and the arteriovenous difference in oxygen content (Ca-vO_2_) based on Fick’s equation [CO=VO_2_/(Ca-vO_2_), where CO is cardiac output which equals SV x HR]. O_2_ pulse at peak of exercise, absolute value or percentage predicted, is a measure of cardiovascular performance on exercise and is prognostic in heart failure (15–18). However, peak O_2_ pulse has not consistently distinguished between patients with and without cardiac dysfunction (19–22). An alternative method is to assess the O_2_ pulse curve over either workload or time rather than its peak value. The curve normally has a hyperbolic profile, with reliance more on SV in the initial and intermediate phases of exercise and more on oxygen extraction and Ca-vO_2_ towards the end until it reaches an asymptotic value (23). Therefore, a shallow or plateaued curve, especially in the early phases of exercise, may represent more cardiovascular than oxygen extraction limitation. A qualitative assessment of the curve trajectory (19, 21), and the estimation of its slope during incremental exercise (24) have been tested in various populations, but not in PH.

The CPET derived slope of ventilation to carbon dioxide production (VE/VCO_2_), which represents the amount of ventilation in L/min required to eliminate 1 L of carbon dioxide, is also associated with cardiac function. Based on Bohr’s equation, VE/VCO_2_ can be elevated when there is either ventilation/perfusion (VQ) mismatch and increased physiological dead-space (V_D_/V_T_) or low arterial partial pressure of carbon dioxide (PaCO_2_) due to hyperventilation (5). In heart failure, an elevated VE/VCO_2_ is caused by both mechanisms; VQ mismatch due to low CO and capillary perfusion, and hyperventilation due to enhanced chemoreceptor sensitivity (25).

The ratio of the VE/VCO_2_ slope to peak VO_2_ (VEVCO_2_/peakVO_2_) integrates both ventilatory efficiency and aerobic capacity. It seems to differentiate between cardiovascular limitations and deconditioning (26) and be a stronger prognostic marker than VE/VCO_2_ slope or peakVO_2_ separately, in patients with left ventricular failure (27).

In this study we sought to examine the role of O_2_ pulse kinetics (curve trajectory and slopes) and VEVCO_2_/peakVO_2_ in identifying cardiovascular limitation in patients with exertional dyspnoea, and mild PAH or CTEPH. Our hypothesis was that in patients with exertional cardiac dysfunction the slopes of O_2_ pulse would be lower (shallower), O_2_ pulse curve would be plateauing earlier, and VEVCO_2_/peakVO_2_ would be higher compared to those without. We also hypothesized that VEVCO_2_/peakVO_2_ ratio would have stronger prognostic value than VE/VCO_2_ slope or peak O_2_ pulse in this population.

## Methods

We included consecutive patients referred to the CPET laboratory in Sheffield Pulmonary Vascular Disease Unit from January 2019 to May 2025. All patients were symptomatic with exertional dyspnoea. Patients were referred for CPET with the following indications: i) unexplained dyspnoea in patients with low probability of PH on echocardiography or no PH on right heart catheterisation (RHC) and risk factors for PAH (e.g. systemic sclerosis), ii) identification of the main cause of dyspnoea in patients with chronic thromboembolic pulmonary disease but no or mild PH, and iii) assessment of the response to targeted therapies/prognostication in patients with PAH and CTEPH. All patients underwent a CPET based on a previously published protocol (Supplementary material).

### RHC and Cardiac Magnetic Resonance Imaging (MRI)

RHC was performed using standard procedures (30) and mean pulmonary artery pressure (mPAP), pulmonary artery wedge pressure (PAWP), CO, cardiac index (CI), and PVR were obtained. CO was measured using the thermodilution method. Some of the patients had also undergone a cardiac MRI prior to CPET and RV ejection fraction was calculated using an established technique with high interobserver reproducibility (31).

### O_2_ pulse slopes and VEVCO_2_/peakVO_2_

We extrapolated raw data of O_2_ pulse obtained every 5 seconds from our CPET system. Slopes were calculated using lines of best fit (y = *α*x + b; *α* is slope, y is O_2_ pulse in ml/beat and x time in minutes) at different phases of the graph. We calculated the following O_2_ pulse slopes: i) slope 1: work phase only (from the end of warm-up phase to the beginning of recovery), ii) slope 2: both warm-up and work phase up to the beginning of recovery, iii) slope 3: recovery phase, iv) slope 4: work phase prior to plateau or decline only in tests exhibiting plateau or decline (Supplementary Figure 1). Plateau of the curve was identified both by inspection of the raw data and visually. Plateau was defined as a less than 0.5 ml/beat increase in O_2_ pulse persisted up to exercise cessation (21). Down-sloping of the curve was defined as a progressive decrement in O_2_ pulse up to exercise cessation lasting for at least 30 sec. Visual assessment was performed by two independent expert CPET interpreters, and their agreement was evaluated. In Qualitative O_2_ pulse analysis, four distinct patterns were identified as previously described (19): 1=up-sloping curve, 2=early plateauing curve (plateau occurs before the 65% of the whole work phase), 3=late plateauing curve (plateau occurs after the 65% of the whole work phase), 4=down-sloping (Supplementary figure 2). If the O_2_ pulse curve was very erratic or consisted of very few datapoints, slopes were not calculated and a pattern was not assigned.

VEVCO_2_/peakVO_2_ was calculated as the ratio of VE/VCO_2_ slope to peak VO_2_ in ml/kg/min (26, 27).

### Group comparisons

We divided patients into three groups: 1) ‘PH’ group: mPAP > 20 mmHg, PAWP ≤ 15 mmHg, and PVR > 2 Wood units on RHC. All patients in this group had either PAH or CTEPH, 2) ‘No PH’ group: mPAP ≤ 20 mmHg or low probability for PH on echocardiogram when RHC had not been performed, and 3) “Unclassified” group: mPAP > 20 mmHg, PAWP ≤ 15 mmHg, and a PVR ≤ 2 Wood units. All the above definitions were based on the current ESC/ERS PH Guidelines (32). We also compared patients’ slopes and Qualitative O_2_ pulse, regardless of PH, according to previously reported cut-off limits of cardiac dysfunction (16–18, 26, 27).

### Statistical methods

Continuous variables were assessed for normality using the Shapiro–Wilk test. Normally distributed variables are presented as mean ± standard deviation. Non-normal data are median and interquartile range (IQR). Categorical variables are summarized as counts and percentages. Comparisons between the three groups (“PH”, “No PH”, “Unclassified”) and the four patterns of Qualitative O_2_ pulse analysis were performed using one-way analysis of variance (ANOVA) for normally distributed continuous variables, or the Kruskal–Wallis test for non-normally distributed ones. For other comparisons independent samples t-test and Wilcoxon rank-sum tests were used for normally and non-normally distributed continuous variables respectively.

Categorical variables were compared using the Chi-square test or Fisher’s exact test when expected cell counts were small. Interobserver variability in identifying the time point of the onset of plateauing or decreasing of the O_2_ pulse curve was assessed using the intraclass correlation coefficient (ICC) and Bland-Altman analysis. Correlation between continuous variables was assessed by using Pearson’s correlation coefficient (r). In our univariable/multivariable logistic regression analysis, peak O_2_ pulse < 65 % predicted was used as the dependent variable as a more robust than < 80% marker of cardiovascular limitation.

Slopes 1 and 3, VEVCO_2_/peakVO_2_, AT % predicted, VO_2_/work, chronotropic index, peak VO_2_ in ml/kg/min, VE/VCO_2_ slope, and Qualitative O_2_ pulse were our independent variables.

Qualitative O_2_ pulse was used as a binary variable (‘normal patterns’= 1 or 3, ‘abnormal patterns’= 2 or 4). Continuous slope variables were scaled per IQR and slope 3 (recovery) being a negative number was inverted before scaling by multiplying by -1 to align the direction of effect. For each continuous variable we calculated optimal cut-off using Youden’s index. In the multivariable analysis we included variables which were statistically significant in the univariable analysis but not VE/VCO_2_ slope or peak VO_2_ to avoid collinearity with VEVCO_2_/peakVO_2_. In all analyses statistical significance was set at p < 0.05. Kaplan-Meier survival analysis was performed to evaluate all-cause mortality stratified by VEVCO_2_/peakVO_2_ < vs. ≥ 2.7 (26), VE/VCO_2_ slope ≥ vs. < 34 (33–35), peak O_2_ pulse % ≥ vs. <65 (18), and slope 1 ≥ vs. < 0.40 (median from this analysis). Time-to-event was defined as the number of days from baseline to death or census date. Patients alive at the census date were censored. To determine which variable was the stronger predictor of survival, we performed a multivariable Cox proportional hazards analysis. Finally, we drew Sankey/alluvial plots to demonstrate the flow of patients with or without PH across the different variables of cardiac dysfunction. All analyses were performed in R version 4.5.2 (R Foundation for Statistical Computing, Vienna, Austria).

### Ethics

Ethical approval was granted by the Institutional Review Board and approved by the National Research Ethics Service (16/YH/0352 subsequently 22/EE/0011). This study was approved by the ASPIRE data management committee and data were extracted and anonymised as per ASPIRE standard operating policy.

## Results

We included 289 out of 333 CPETs in our analysis. We excluded 44 repeat tests to analyse only one test, the closest to RHC, per patient. In 6 tests (2%) we were unable to calculate VEVCO_2_/peakVO_2_ as VE/VCO_2_ trace was too disorganised due to dysfunctional breathing. In 40 tests (14%) we were unable to estimate all the slopes for the same reason. A hundred and eighty-one (181) patients had had a cardiac MRI prior to CPET (62.6%). On average, tests were maximal (RER > 1.1 and peak heart rate > 80% predicted) and median haemoglobin normal across all groups [155 (144-168) g/L]. The agreement in the Qualitative O_2_ pulse analysis between the two operators was good to excellent (ICC=0.899. 95% CI 0.732-0.955, p < 0.001) and Bland-Altman analysis showed an average bias of 0.5 minutes (Supplementary figure 3).

One hundred and thirty-six (136) patients had pre-capillary PH, 46 “unclassified” PH, and 107 no PH. In the “no PH” group 39 (36%) patients had RHC data. Demographics of the groups are presented in Supplementary table 1 and CPET parameters in Table 1. The slopes were not different between the groups, but Qualitative O_2_ pulse patterns differed with a higher proportion of pre-capillary PH patients having patterns 2-4. Peak VO_2_, AT, peak O_2_ pulse % predicted and indices of ventilatory efficiency were more impaired in the pre-capillary PH group.

**Table 1.**
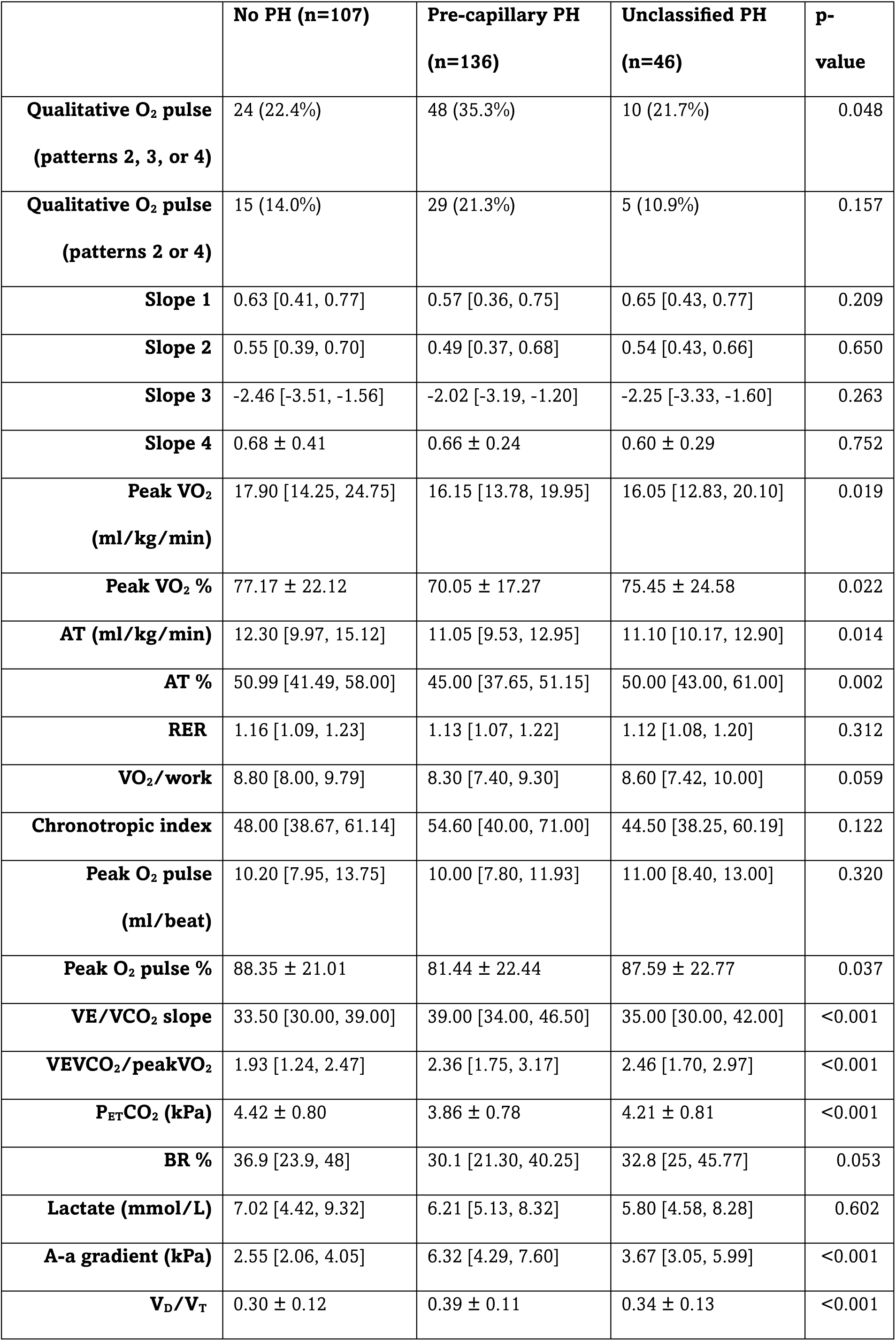
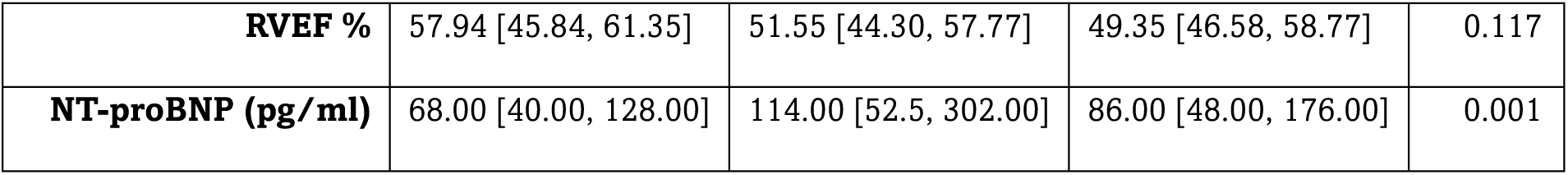
CPET, MRI, NT-proBNP data in patients with and without PH. A-a gradient, alveolar-arterial oxygen difference; AT, anaerobic threshold; BR, breathing reserve; NT-proBNP, N-terminal brain natriuretic peptide; O_2_, oxygen; P_ET_CO_2_, partial pressure of end-tidal carbon dioxide; PH, pulmonary hypertension; RER, respiratory exchange ratio; RVEF, right ventricular ejection fraction; Slope 1, slope of O_2_ pulse with no warm-up phase; slope 2, slope of both warm-up and work phase; slope 3, slope of O_2_ pulse in recovery; slope 4, slope of O_2_ pulse before the plateau; VE/VCO_2_, slope of ventilation to carbon dioxide production; V_D_/V_T_, physiological dead-space as a ratio to tidal volume; VO_2_, oxygen uptake, VO_2_/work, oxygen uptake per unit of work.

After dividing patients into the 4 different patterns of Qualitative O_2_ pulse, slopes 1, 2 and 3 were significantly lower (shallower), peak O_2_ pulse lower and chronotropic index higher (steeper HR response) in patterns 2 and 4 compared to 1 and 3 (Table 2). When all patients were divided by peak O_2_ pulse % predicted (≥ or < 80% and ≥ or < 65%) and VEVCO_2_/peakVO_2_ (< or ≥ 2.7), all slopes were lower and all O_2_ delivery parameters more impaired in the < 80%, <65% and ≥ 2.7 groups (Supplementary tables 2, 3, 4). Finally, we identified slope < 0.40 as the cut-off limit of possible cardiac dysfunction for slope 1 by its median value in patients with VEVCO_2_/peakVO_2_ ≥ 2.7 and peak O_2_ pulse < 80%. After dividing patients into < or ≥ 0.40, those with shallower slopes had more impaired O_2_ delivery parameters indicating cardiac dysfunction (Supplementary table 5 and supplementary figure 4). A moderate but statistically significant correlation between peak O_2_ pulse % predicted, VEVCO_2_/peakVO_2_ and all the slopes was shown (Supplementary figures 5 and 6). The correlation between peak VO_2_ and VE/VCO_2_ slope was weak (Supplementary figure 7).

**Table 2.**
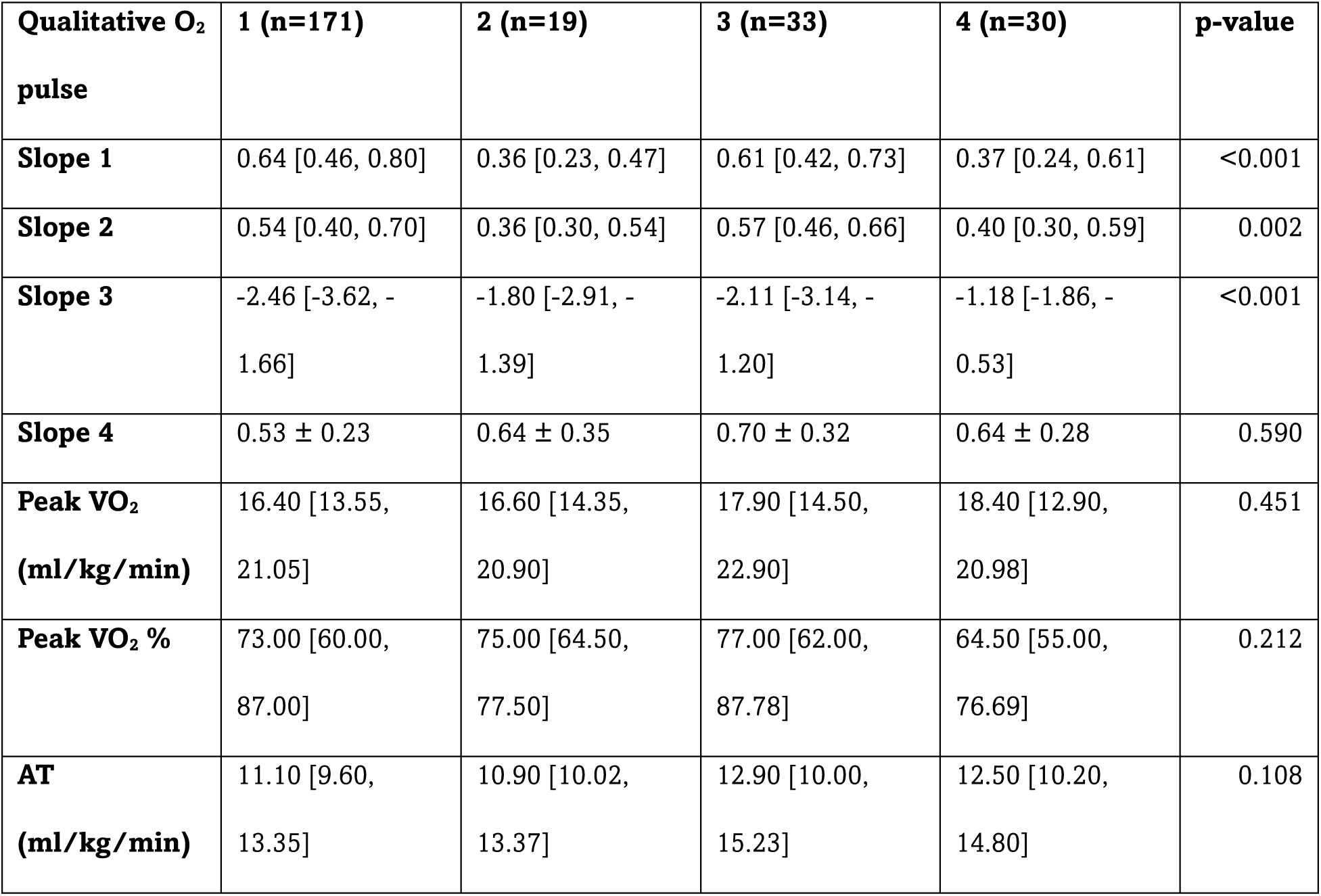
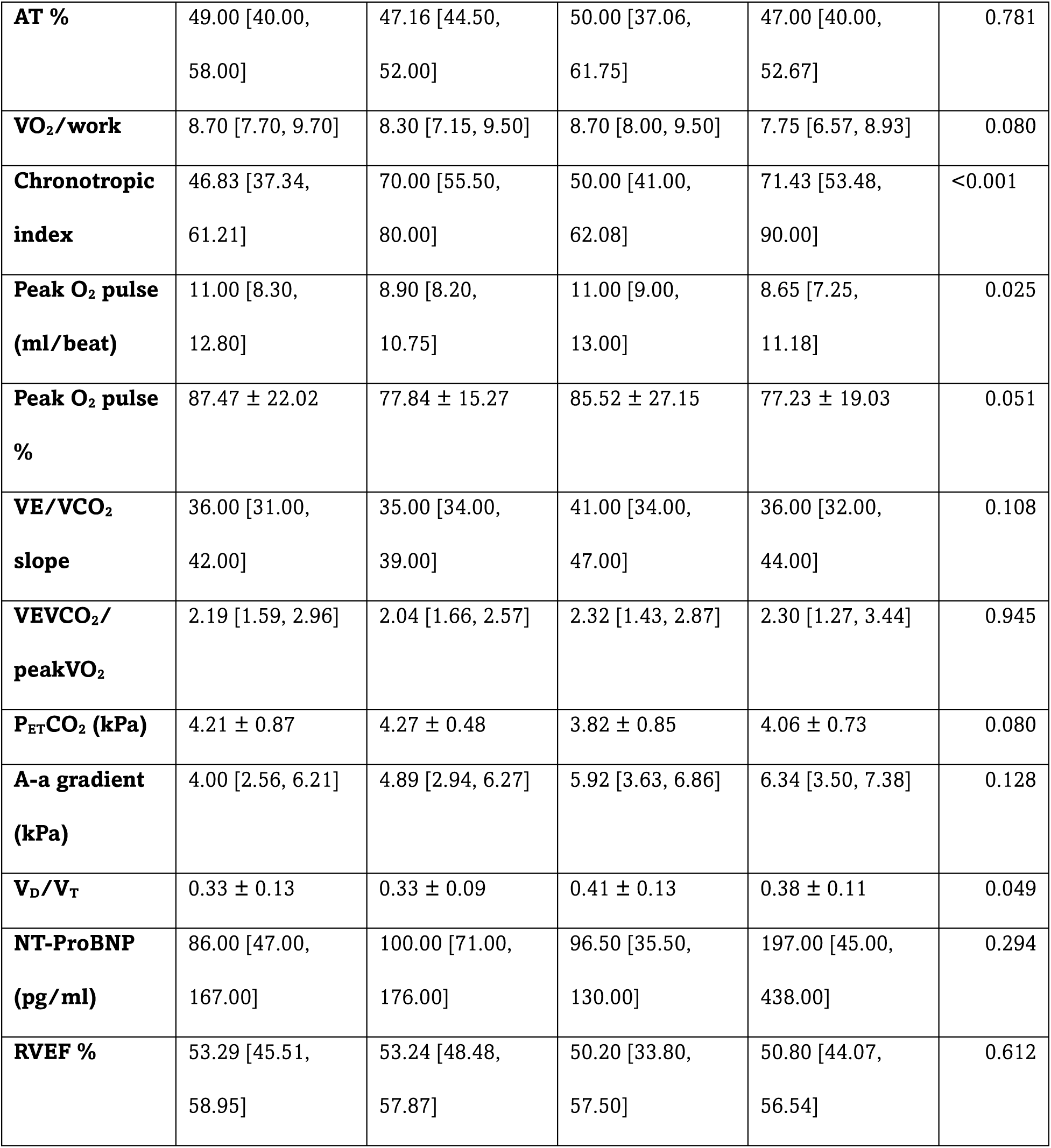
CPET parameters according to Qualitative O_2_ pulse patterns. A-a gradient, alveolar-arterial oxygen difference; AT, anaerobic threshold; BR, breathing reserve; NT-proBNP, N-terminal brain natriuretic peptide; O_2_, oxygen; P_ET_CO_2_, partial pressure of end-tidal carbon dioxide; RER, respiratory exchange ratio; RVEF, right ventricular ejection fraction; Slope 1, slope of O_2_ pulse with no warm-up phase; slope 2, slope of both warm-up and work phase; slope 3, slope of O_2_ pulse in recovery; slope 4, slope of O_2_ pulse before the plateau; VE/VCO_2_, slope of ventilation to carbon dioxide production; V_D_/V_T_, physiological dead-space as a ratio to tidal volume; VO_2_, oxygen uptake, VO_2_/work, oxygen uptake per unit of work.

The univariable/multivariable analysis, forest plots and ROC curves are shown in table 3, supplementary table 6, and supplementary figures 8 and 9 respectively. VEVCO_2_/peakVO_2_ increased the risk of cardiac dysfunction by 3.9 times and VE/VCO_2_ slope by 2.6, although the latter had lower sensitivity than VEVCO_2_/peakVO_2_ (0.72 vs. 0.86). The optimal cut-off limit for VEVCO_2_/peakVO_2_ identified in our analysis was 2.4 (AUC 0.83). In the multivariable analysis only VEVCO_2_/peakVO_2_ and AT % retained statistical significance.

**Table 3.** Univariable analysis. AT, anaerobic threshold; slope 1, slope of O_2_ pulse with no warm-up phase; slope 3, slope of O_2_ pulse in recovery; VE/VCO_2_, slope of ventilation to carbon dioxide production; VO_2_, oxygen uptake, VO_2_/work, oxygen uptake per unit of work.

| Variable | OR (95% CI) | p-value | AUC | Optimal cut-off | Sensitivity | Specificity |
| --- | --- | --- | --- | --- | --- | --- |
| **Slope 1** | 0.16 (0.08-0.29) | <0.001 | 0.81 | 0.36 | 0.69 | 0.88 |
| **Slope 3** | 0.14 (0.06-0.29) | <0.001 | 0.80 | 1.17 | 0.59 | 0.87 |
| **VEVCO<sub>2</sub>/peakVO<sub>2</sub>** | 3.89 (2.60-6.17) | <0.001 | 0.83 | 2.37 | 0.86 | 0.65 |
| **VE/VCO<sub>2</sub> slope** | 2.61 (1.80-3.89) | <0.001 | 0.73 | 38.5 | 0.72 | 0.66 |
| **Peak VO<sub>2</sub>**<br>**(ml/kg/min)** | 0.10 (0.04-0.22) | <0.001 | 0.82 | 13.2 | 0.67 | 0.87 |
| **AT %** | 0.28 (0.15- 0.48) | <0.001 | 0.83 | 41.2 | 0.71 | 0.78 |
| **VO<sub>2</sub>/work** | 0.28 (0.18-0.44) | <0.001 | 0.76 | 7.94 | 0.67 | 0.74 |
| **Chronotropic index** | 3.30 (2.25-5.00) | <0.001 | 0.77 | 62.1 | 0.69 | 0.77 |
| **Qualitative O<sub>2</sub> pulse** | 1.82 (0.81-3.89) | 0.113 | 0.55 | - | 0.28 | 0.82 |

In our survival analysis, in 193 patients with VEVCO_2_/peakVO_2_ < 2.7 and median time to census 1287 days, there were 3 deaths, whilst in 90 patients with ≥ 2.7 (968 days) 15. Kaplan-Meier curves demonstrated significantly lower survival in the VEVCO_2_/peakVO_2_ ≥ 2.7 group (log-rank p < 0.001). Sixty-two (62) patients had slope 1 < 0.40 (839 days) and 187 ≥ 0.40 (1210 days) with 5 deaths in the < 0.40 group and 11 in the ≥ 0.40 group (log-rank p = 0.20). In the peak O_2_ pulse < 65% (n = 45) median follow-up was 874 days, and there were 6 deaths, whilst in the ≥ 65% (n = 244), 1245 days and 13 deaths. There was a significantly lower survival rate in patients with < 65% compared with those ≥ 65% (log-rank p =0.014). Finally, in the VE/VCO_2_ < 34 group (n=101), median follow up was 1414 days and there was one death, whilst in the ≥ 34 (n=182) it was 1120 days with 17 deaths respectively (p=0.002) (Figure 1). After adjustment for age, sex, mPAP, RAP and PVR, peak O_2_ pulse < 65% and VE/VCO_2_ lost statistical significance (HR 0.93, 95% CI 0.32–2.69, p = 0.897 and HR 3.61, 95% CI: 0.42–31.1, p = 0.243 respectively), whereas VEVCO_2_/peakVO_2_ ≥ 2.7 remained a strong independent predictor of mortality (HR 13.6, 95% CI 3.8–48.5, p < 0.001).

**Figure 1.**
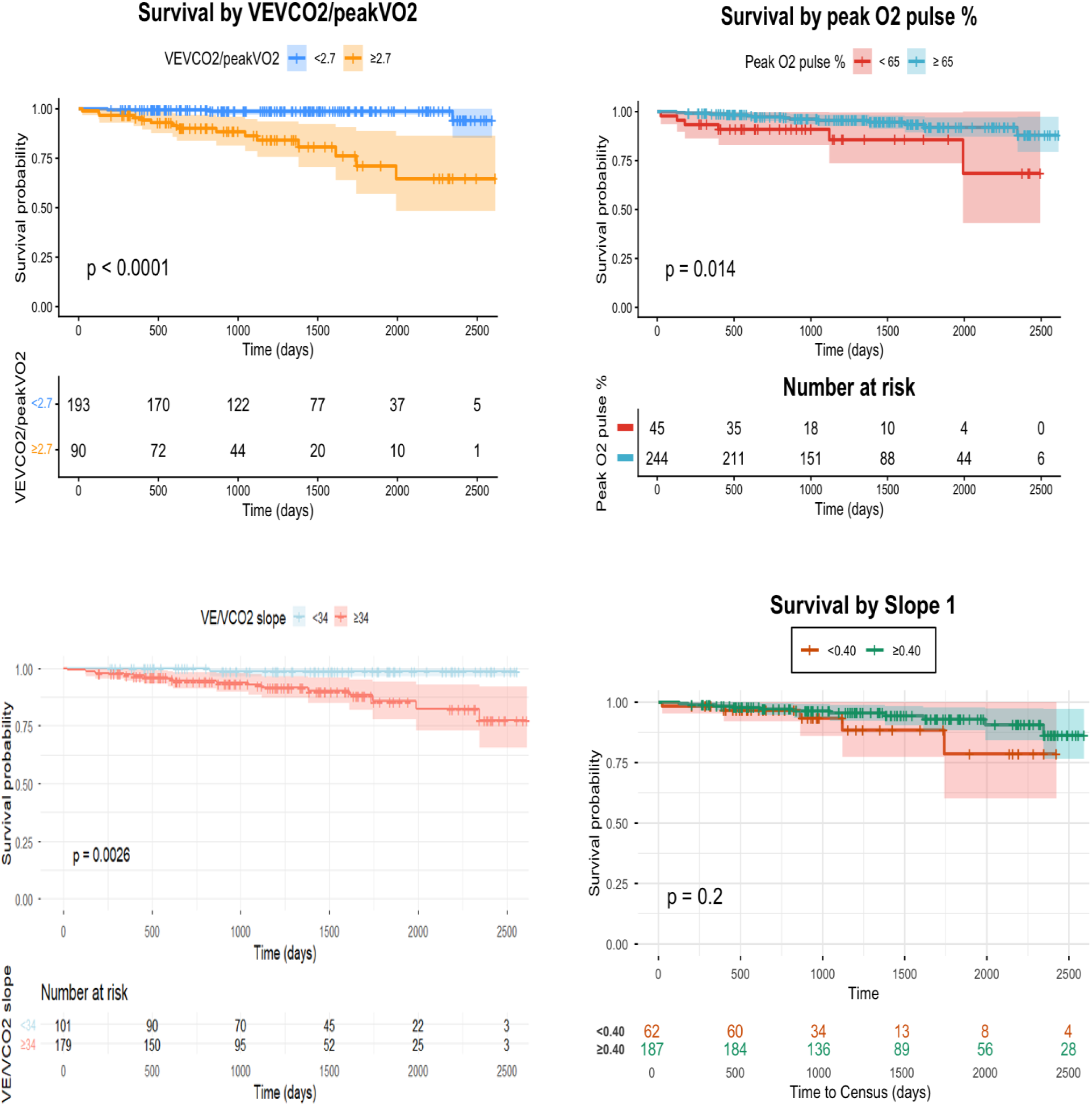
Kaplan-Meier curves. O_2_, oxygen; slope 1, slope of O_2_ pulse with no warm-up phase; VE/VCO_2_, slope of ventilation to carbon dioxide production; VO_2_, oxygen uptake.

In the Sankey/alluvial graphs patients with pre-capillary PH appeared to have more impaired markers of cardiac dysfunction than without, whilst approximately 2/3 of the patients with “unclassified PH” and peak O_2_ pulse < 80% had VEVCO_2_/peakVO_2_ ≥ 2.7 (Figure 2 and Supplementary figure 10).

**Figure 2.**
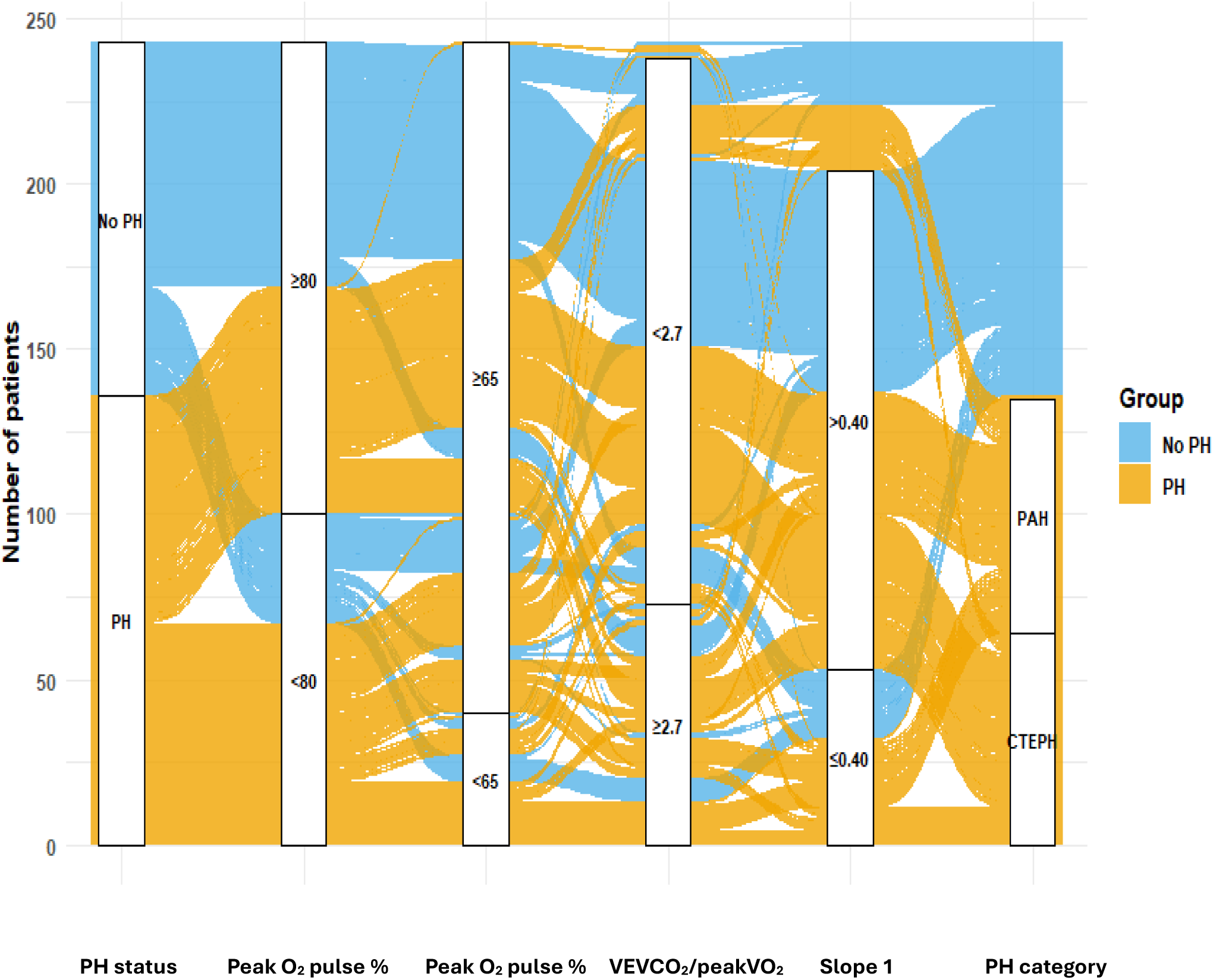
Sankey/alluvial graph showing the distribution of patients with and without pulmonary hypertension based on different markers of cardiac dysfunction. O_2_, oxygen; slope 1, slope of O_2_ pulse with no warm-up phase; VE/VCO_2_, slope of ventilation to carbon dioxide production; VO_2_, oxygen uptake.

## Discussion

The main findings of our study are: i) in patients with mild pre-capillary PH, there is more cardiovascular limitation on exercise than in those without, ii) plateauing or decreasing of O_2_ pulse slope is more frequent in patients with pre-capillary PH than without and is also associated with shallower O_2_ pulse slopes and slower recovery, iii) the slope of O_2_ pulse during the work phase of exercise is a good discriminator of cardiac dysfunction at a cut-off limit of 0.40, iv) the VEVCO_2_/peakVO_2_ ratio was an independent predictor of cardiac dysfunction and a stronger than peak O_2_ pulse % or VE/VCO_2_ predictor of mortality in our cohort.

Degani-Costa et al (21) examined O_2_ pulse trajectory in two completely different groups of patients; in PAH where there is typically low SV and in mitochondrial myopathy (MM) where there is poor oxygen muscle extraction. Abnormal O_2_ pulse trajectory (“flattened” or “decreasing”) was observed in 67% of the PAH and only in 20% of MM patients, in contrast to peak O_2_ pulse % predicted, VO_2_/work or AT which were equally diminished between the groups. Mapelli et al (19), in patients with hypertrophic cardiomyopathy, showed that abnormal O_2_ pulse trajectory was able to identify patients with compromised oxygen delivery, whilst left ventricular outflow tract gradient did not. In our study, flattened or down-sloping O_2_ pulse curves (patterns 2, 3, and 4) were observed more often in the pre-capillary PH compared to the non-PH group, but we did not show a difference in “abnormal” patterns (2 or 4) versus the “normal” ones (1 or 3) possibly due to the relatively modest haemodynamics of our pre-capillary PH cohort. In addition, we showed that patients with early plateauing and down-slopping curves had more impaired CPET markers of cardiac dysfunction. This observation is consistent with known pathophysiology in heart failure. In patients with heart failure, although SV increase is blunted on exercise, peripheral oxygen use is not. Thus, Ca-vO_2_ widens at the beginning of exercise and remains reasonably preserved throughout via optimised peripheral blood flow redistribution and high mitochondrial activity (36, 37). Therefore, inadequate SV increase becomes the predominant, although not the sole, cause of exercise limitation and a shallow or plateaued curve represents pump failure than oxygen extraction limitations.

The slope of the O_2_ pulse curve during incremental exercise has been calculated previously in patients with a Fontan circulation and it showed to outperform peak O_2_ pulse as a surrogate marker of SV on exercise (24). In our study, we expanded O_2_ pulse slope calculation to recovery and to the slope before the plateauing or decline of the curve. Patients with lower peak O_2_ pulse % (based on two different cut-off limits) had significantly lower O_2_ pulse slopes and slower recovery compared to the higher ones. We also identified a cut-off limit for slope 1 at 0.40 and we showed that when the slope was < 0.40, all CPET parameters indicating cardiac dysfunction were significantly worse than in ≥ 0.40. Slopes were not independent predictors of cardiac limitation in our multivariable analysis, and they did not predict survival in our cohort. One possible explanation is that O_2_ pulse curves are not always linear and a regression line may not perfectly fit to represent the true course of the curve. Another possible explanation may be the absence of severe cardiac dysfunction in our cohort. In our logistic regression and survival analyses, we selected slope 1 (warm-up phase excluded) instead of 2 (whole line) as we noticed that during the warm-up phase O_2_ pulse curve often plateaus quickly affecting the linearity of the whole curve.

Both peak VO_2_ and VE/VCO_2_ have prognostic value in heart failure (38–41), however, some patients with heart failure have normal peak VO_2_, whist VE/VCO_2_ remains high maintaining its ability to predict outcome (42). This observation indicates that changes in these two parameters in heart failure do not necessarily reflect the same pathophysiology. Consistent with this observation is the weak correlation between peak VO_2_ and VEVCO_2_ in our study. Guazzi et al (27) suggested indexing VE/VCO_2_ slope by peak VO_2_ and showed that a VEVCO_2_/peakVO_2_ ≥ 2.4 was a stronger predictor of survival compared to VE/VCO_2_ and peak VO_2_ alone in patients with ischaemic and dilated cardiomyopathy. Rosenbaum et al (26) used VEVCO_2_/peak VO_2_ to discriminate patients with heart failure from those with deconditioning by performing both CPET and stress echocardiography. Stroke volume index <50 ml/m^2^ on exercise was identified as a “gold-standard” of cardiovascular limitation and a VEVCO_2_/peak VO_2_ ≥ 2.7 was suggestive of cardiac failure in that study. In our study VEVCO_2_/peakVO_2_ was an independent predictor of cardiac failure, along with AT%, and a strong predictor of mortality. It is interesting that the optimal cut-off limit for VEVCO_2_/peak VO_2_ that we observed was 2.4, exactly as in the study by Guazzi et al (27). In patients with poor oxygen muscle extraction such as in deconditioning, VE/VCO_2_ is expected to be normal, unless if there is concomitant hyperventilation for other reasons (e.g. dysfunctional breathing). Thus, a high VEVCO_2_/peak VO_2_ as a discriminator between patients with cardiac dysfunction and deconditioning seems to make physiological sense. In addition, in a population with PAH and CTEPH, where VE/VCO_2_ can be elevated due to chronic clots or pulmonary artery remodelling rather than cardiac failure, indexing ventilatory inefficiency with peak VO_2_ may be helpful to discriminate between the two.

This study has the limitations of a retrospective single-centre study. The selected cohort was heterogeneous with regards to the indication for the exercise test, and not all patients in the non-PH group had PH excluded by RHC. In addition, a possibly modest severity of cardiac dysfunction in our population may have affected our power to detect significant differences in some of the comparisons that we made. The greatest challenge has been the absence of a “gold-standard” metric of cardiac failure on exercise. This should be a metric showing the inability of the heart to adequately increase SV to meet its metabolic demands on exercise. This could possibly be SV index, although a cut-off limit for it is far from established. In our cohort we lacked exercise haemodynamics and therefore a non-CPET benchmark metric for cardiovascular limitation. The absence of a “gold-standard” did not allow us to examine the additional value of the slopes and VEVCO_2_/peakVO_2_ ratio to the current established CPET oxygen delivery parameters. This need should be addressed in future research.

## Conclusion

In a cohort of patients with and without PH, VEVCO_2_/peakVO_2_ was an independent predictor of cardiovascular limitation on exercise and a strong predictor of mortality. This metric may be particularly useful in patients with mild pre-capillary PH to discriminate between cardiac failure and deconditioning, where other CPET parameters of oxygen delivery may not be able to.

## Data Availability

All data produced in the present study are available upon reasonable request to the authors

