## Supplementary material for "Oxygen pulse kinetics and ventilatory inefficiency as markers of cardiovascular limitation on exercise in patients with mild pre-capillary pulmonary hypertension and exertional dyspnoea"

**CPET protocol**

Patients underwent CPET on an upright cycle ergometer using a breath-by-breath system (Vyntus CPX, Vyaire Medical, Germany) according to the American Thoracic Society/American Society of Chest Physicians statement on CPET (28). Each test was performed in three stages: 3 minutes of rest, 3 minutes of unloaded pedaling (warm-up phase), and a progressive ramp increase in workload to maximum exercise, with the ramp rate estimated to result in 8 to 12 minutes of exercise (work phase). Systemic blood pressure was measured with a sphygmomanometer, finger or earlobe oxygen saturation was measured with a pulse oximeter, and a 12-lead electrocardiogram was continuously recorded. Face mask breath-by-breath measures included oxygen uptake ($VO_2$), carbon dioxide production ($VCO_2$), and ventilation (VE). An arterial blood sample was drawn just after termination of exercise for measurement of arterial partial pressure of oxygen ($PaO_2$), arterial partial pressure of carbon dioxide ($PaCO_2$) and lactate level. Peak $VO_2$ was averaged over the last 30 seconds of exercise. The best estimate of anaerobic threshold (AT) was calculated manually using a combination of the V-slope method and ventilatory equivalents for oxygen. $VE/VCO_2$ slope was obtained by linear regression analysis of the relationship between VE and $VCO_2$ during exercise prior to the respiratory compensation point. Breathing reserve (maximal voluntary ventilation – peak ventilation) was calculated as 40 x FEV1. Spirometry was performed prior to each test. Chronotropic index was calculated as: (peak heart rate-resting heart rate) / (peak $VO_2$ in ml/L-resting $VO_2$ in ml/L) (29). Physiological dead space ($V_D/V_T$) was measured using the Bohr equation: $V_D/V_T = (PaCO_2 - P_ECO_2)/PaCO_2$, where $P_ECO_2$ is the measured mixed expired partial pressure of $CO_2$. $VO_2$, $VCO_2$, VE, heart rate and all their derivative metrics were also measured and recorded for 2 minutes on average in recovery.

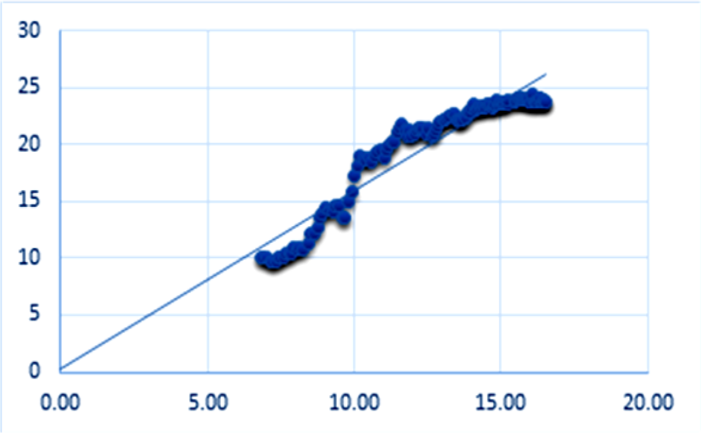
30
25
20
15
10
5
0
0.00
5.00
10.00
15.00
20.00

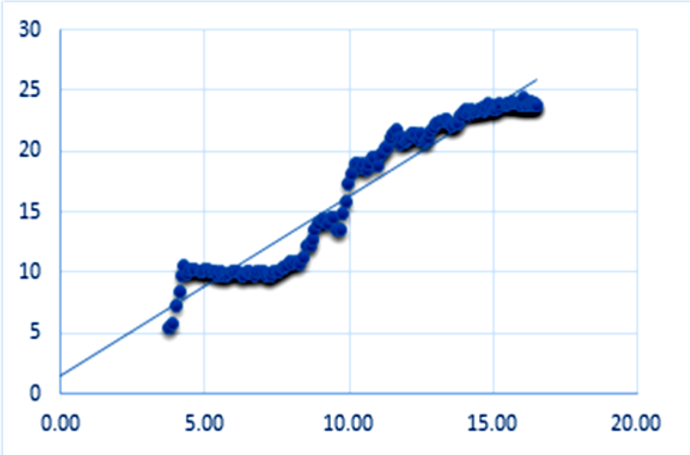
30
25
20
15
10
5
0
0.00
5.00
10.00
15.00
20.00

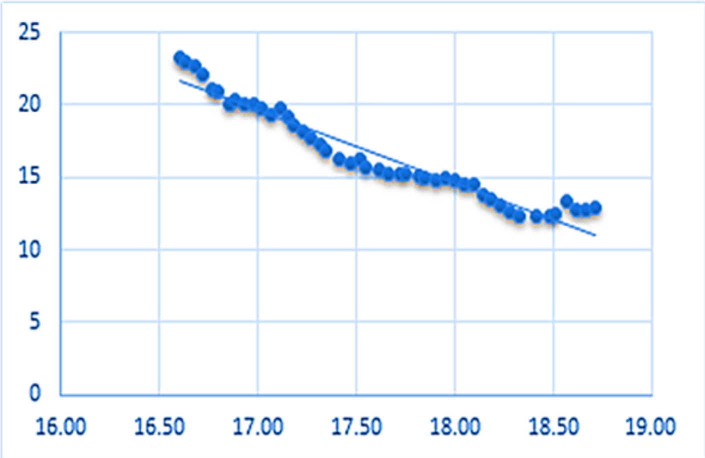
25
20
15
10
5
0
16.00
16.50
17.00
17.50
18.00
18.50
19.00

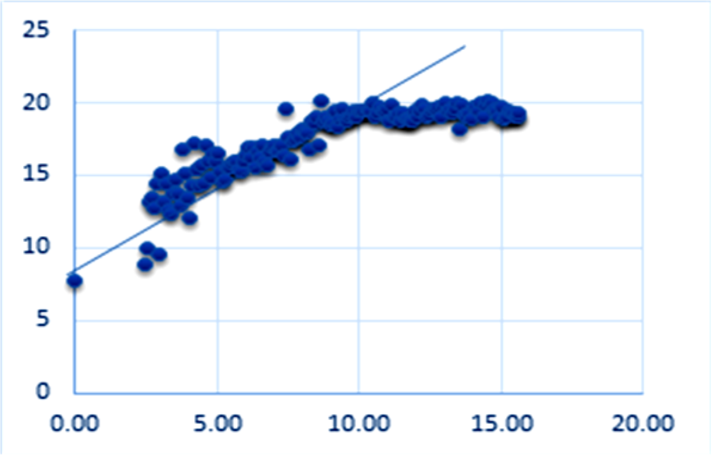
25
20
15
10
5
0
0.00
5.00
10.00
15.00
20.00

**Supplementary figure 1. Slopes of $O_2$ pulse curves**.

*From top to bottom: slope 1: work phase only (from end of warm-up to peak exercise, excluding warm-up), slope 2: entire exercise phase (including warm-up to peak exercise), same patient with slope 1, slope 3: recovery phase (first 2 minutes post-exercise), slope 4: work phase prior to plateau or decline (from end of warm-up to the visually identified inflection point where the $O_2$ pulse–time relationship plateaued or decrease).*

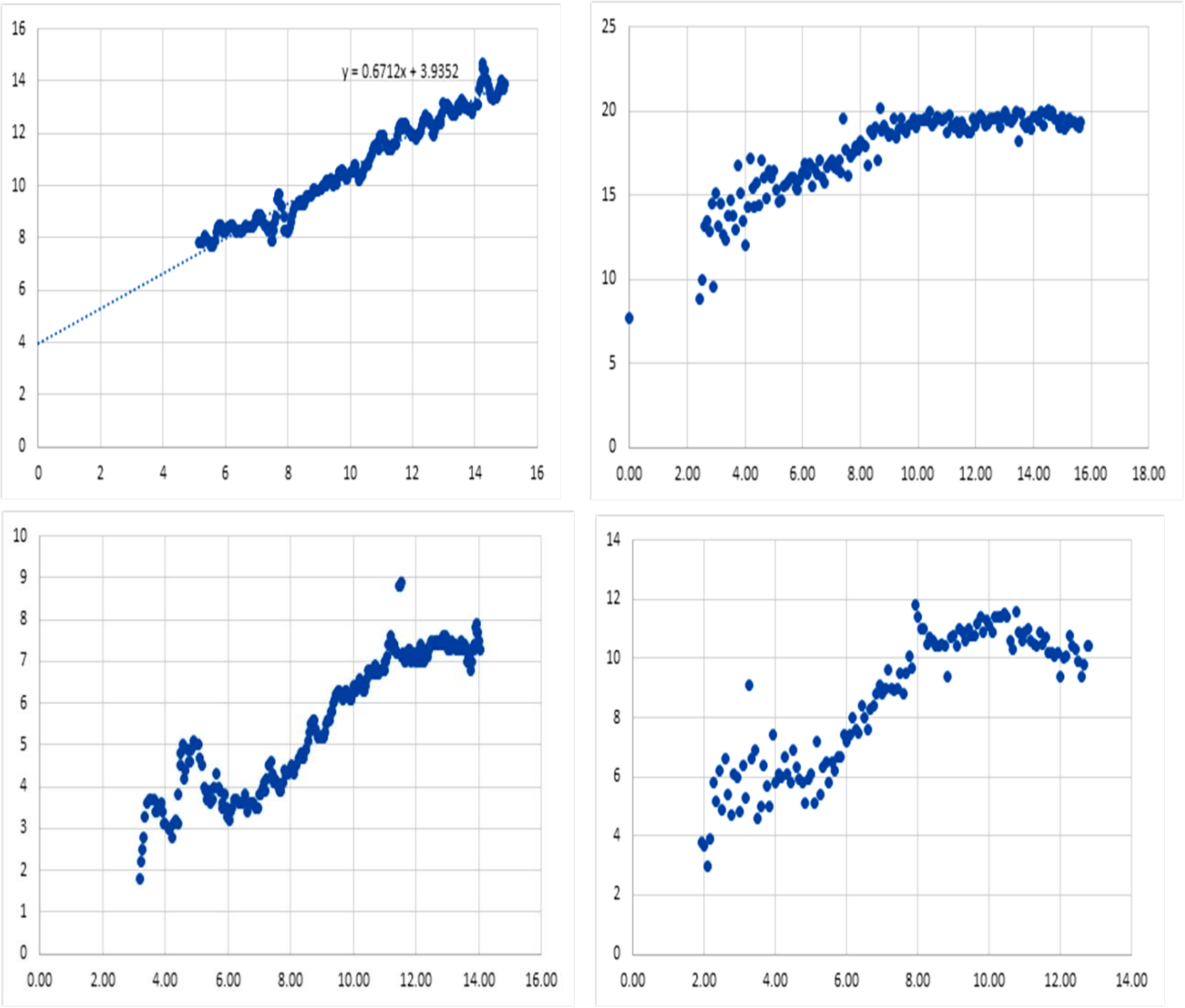


**Supplementary figure 2. Qualitative $O_2$ pulse patterns**.

*On y-axis it is $O_2$ pulse in ml/beat and on x-axis time in minutes. Top left: pattern 1, up-sloping $O_2$ pulse curve, top right: pattern 2, early plateauing, bottom right: pattern 3, late plateauing, and bottom right: pattern 4, down-sloping. In the top left panel, a line of best fit and the equation for the slope is shown.*

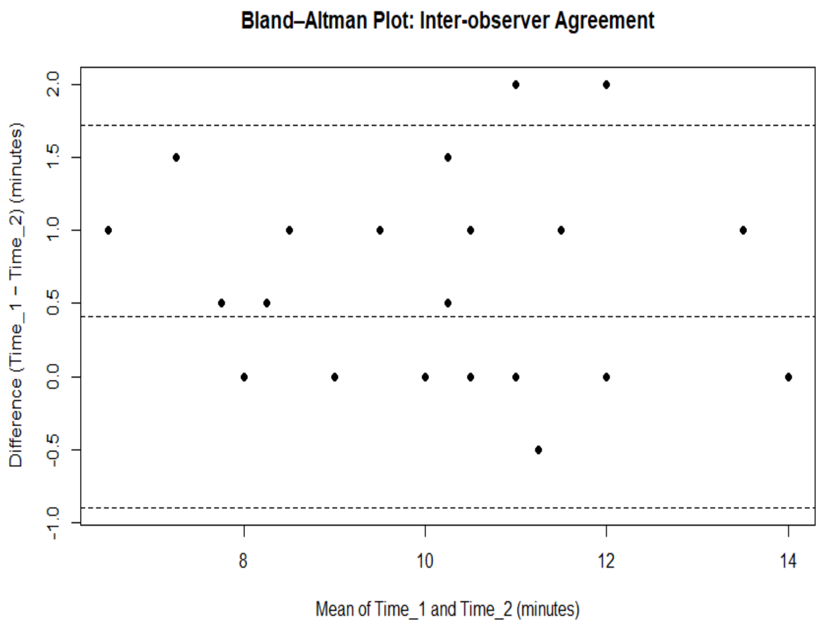


**Supplementary figure 3. Bland-Altman analysis**

*Inter-observer variability in the qualitative assessment of $O_2$ pulse trajectory (time to plateau or drop).*

| | No PH (n=107) | Pre-capillary PH (n=136) | Unclassified PH (n=46) | p-value |
|---|---|---|---|---|
| **Age (years)** | 55.00 [38.00, 64.50] | 53.50 [41.00, 66.00] | 53.50 [47.00, 63.25] | 0.792 |
| **Male (%)** | 45 (42.1) | 73 (53.7) | 24 (52.2) | 0.179 |
| **BMI (kg/m$^2$)** | 28.27 [24.71, 32.24] | 28.07 [24.95, 33.72] | 31.61 [27.50, 34.39] | 0.010 |
| **Systemic hypertension (%)** | 25 (23%) | 37 (27%) | 12 (26%) | 0.799 |
| **Atrial fibrilation (%)** | 4 (3.7%) | 8 (5.8%) | 2 (4.3%) | 0.816 |
| **Diabetes mellitus (%)** | 6 (5.6%) | 18 (13.2%) | 7 (15%) | 0.073 |
| **COPD (%)** | 10 (9.3%) | 11 (8%) | 4 (8.6%) | 0.959 |
| **Interstitial lung disease (%)** | 5 (4.6%) | 10 (7.3%) | 4 (8.6%) | 0.530 |
| **Ischaemic heart disease (%)** | 5 (4.6%) | 12 (8.8%) | 3 (2.2%) | 0.450 |
| **FEV1 (L)** | 2.75 [2.12, 3.37] | 2.54 [1.97, 3.12] | 2.70 [2.20, 3.09] | 0.337 |
| **FEV1 %** | 89.60 [72.55, 102.40] | 82.50 [71.00, 93.85] | 85.00 [78.00, 96.25] | 0.057 |
| **FVC (L)** | 3.52 [2.81, 4.39] | 3.58 [2.76, 4.24] | 3.64 [3.13, 4.17] | 0.952 |
| **FVC %** | 92.00 [80.00, 106.00] | 91.00 [79.62, 100.75] | 91.00 [80.75, 100.25] | 0.511 |
| **FEV1/FVC** | 76.00 [71.00, 82.00] | 73.40 [68.00, 78.40] | 78.00 [70.00, 82.34] | 0.020 |
| **RAP (mmHg)** | 4 [2, 6] | 6 [4, 8] | 7 [5, 9] | <0.001 |
| **mPAP (mmHg)** | 18.00 [16.00, 19.00] | 31.00 [26.00, 43.25] | 22.00 [22.00, 25.00] | <0.001 |
| **PAWP (mmHg)** | 8 [6, 11.5] | 10 [8, 12] | 13 [11,15.7] | <0.001 |

| | | | | |
|---|---|---|---|---|
| **Cardiac output (L/min)** | 5.93 [5.42, 6.57] | 5.52 [4.51, 6.46] | 6.57 [5.80, 7.20] | <0.001 |
| **Cardiac index (L/min/m$^2$)** | 3.14 [2.67, 3.65] | 2.85 [2.42, 3.41] | 3.23 [2.93, 3.72] | 0.005 |
| **PVR (dynes)** | 114.00 [91.00, 146.00] | 318.50 [218.00, 519.25] | 121.50 [108.00, 138.00] | <0.001 |

**Supplementary table 1. Demographics of patients with and without PH.**

*BMI, body mass index; COPD, chronic obstructive pulmonary disease, FEV1, forced expiratory volume in 1 second; FVC, forced vital capacity; mPAP, mean pulmonary artery pressure; PAWP, pulmonary artery wedge pressure; PH, pulmonary hypertension; PVR, pulmonary vascular resistance; RAP, right atrial pressure.*

| **Peak $O_2$ pulse %** | **≥ 80 (n=171)** | **< 80 (n=118)** | **p-value** |
|---|---|---|---|
| **Slope 1** | 0.69 ± 0.25 | 0.44 ± 0.23 | <0.001 |
| **Slope 2** | 0.61 ± 0.21 | 0.4 ± 0.18 | <0.001 |
| **Slope 3** | -2.48 [-3.63, -1.62] | -1.81 [-2.74, 0.95] | 0.002 |
| **Slope 4** | 0.78 ± 0.26 | 0.58 ± 0.25 | 0.003 |
| **AT (ml/kg/min)** | 11.5 [10.2-13.53] | 10.15 [9.02-12] | <0.001 |
| **AT %** | 51 [42.59-60] | 42.5 [36.22-49] | <0.001 |
| **$VO_2$/work** | 8.78 ± 1.42 | 7.8 ± 1.61 | <0.001 |
| **Chronotropic Index** | 44 [36-55] | 69 [49.5-88] | <0.001 |
| **Peak $O_2$ pulse (ml/beat)** | 11.75 [9.5-14.77] | 7.7 [6.5-10.5] | <0.001 |
| **Peak $VO_2$ (ml/kg/min)** | 18.5 [14.8, 23.8] | 14.9 [11.9, 18.1] | <0.001 |
| **Peak $VO_2$ % predicted** | 83 [72.5, 95] | 59.5 [51.1, 68] | <0.001 |

| | | | |
|---|---|---|---|
| **VE/VCO$_2$ slope** | 34 [30, 39] | 40 [34, 47] | <0.001 |
| **VEVCO$_2$/peakVO$_2$** | 1.92 [1.27, 2.46] | 2.7 [2.06, 3.73] | <0.001 |

**Supplementary table 2. CPET parameters according to peak O$_2$ ≥ and < 80%.**

*AT, anaerobic threshold; O$_2$, oxygen; slope 1, slope of O$_2$ pulse with no warm-up phase; slope 2, slope of both warm-up and work phase; slope 3, slope of O$_2$ pulse in recovery; slope 4, slope of O$_2$ pulse before the plateau; VE/VCO$_2$, slope of ventilation to carbon dioxide production; VO$_2$, oxygen uptake, VO$_2$/work, oxygen uptake per unit of work.*

| **Peak O$_2$ pulse %** | **≥ 65 (n=244)** | **< 65 (n=45)** | **p-value** |
|---|---|---|---|
| **Slope 1** | 0.63 [0.46, 0.78] | 0.3 [0.23, 0.49] | <0.001 |
| **Slope 2** | 0.56 [0.41, 0.7] | 0.3 (0.22-0.43) | <0.001 |
| **Slope 3** | -2.44 [-3.57, -1.6] | -0.91 [-1.86, -0.44] | <0.001 |
| **Slope 4** | 0.72 ± 0.3 | 0.43 ± 0.2 | <0.001 |
| **AT (ml/kg/min)** | 11.8 [10.2-14.25] | 10 [8.4-10.9] | <0.001 |
| **AT %** | 49.72 [42.12-57.9] | 35 (33-43) | <0.001 |
| **VO$_2$/work** | 8.7 [7.9-9.7] | 7.3 [6.03-8.28] | <0.001 |
| **Chronotropic Index** | 47 [38-61] | 71 [56-98] | <0.001 |
| **Peak O$_2$ pulse (ml/beat)** | 11 [8.5-13.05] | 6.8 [5.3-9] | <0.001 |
| **Peak VO$_2$ (ml/kg/min)** | 17.4 [14.5, 22.1] | 12.6 [10.5, 14.6] | <0.001 |
| **Peak VO$_2$ % predicted** | 76 [64, 88.1] | 51 [44, 55] | <0.001 |
| **VE/VCO$_2$ slope** | 35 [31, 41] | 44 [36, 53.5] | <0.001 |
| **VEVCO$_2$/peakVO$_2$** | 2.06 [1.43, 2.69] | 3.65 [2.46, 4.67] | <0.001 |

**Supplementary table 3. CPET parameters according to peak $O_2$ pulse ≥ and < 65%.**

*AT, anaerobic threshold; $O_2$, oxygen; slope 1, slope of $O_2$ pulse with no warm-up phase; slope 2, slope of both warm-up and work phase; slope 3, slope of $O_2$ pulse in recovery; slope 4, slope of $O_2$ pulse before the plateau; $VE/VCO_2$, slope of ventilation to carbon dioxide production; $VO_2$, oxygen uptake, $VO_2$/work, oxygen uptake per unit of work.*

| **$VEVCO_2/PeakVO_2$** | **< 2.7 (n=193)** | **≥ 2.7 (n=90)** | **p-value** |
|---|---|---|---|
| **Qualitative $O_2$ Pulse (patterns 2 or 4)** | 32 (16 %) | 17 (19%) | 0.611 |
| **Slope 1** | 0.67 [0.54, 0.80] | 0.40 [0.26, 0.58] | <0.001 |
| **Slope 2** | 0.59 [0.46, 0.72] | 0.37 [0.27, 0.47] | <0.001 |
| **Slope 3** | -2.76 [-3.97, -1.84] | -1.22 [-2.02, -0.59] | <0.001 |
| **Slope 4** | 0.78 [0.61, 0.95] | 0.47 [0.33, 0.61] | <0.001 |
| **Peak $VO_2$ (ml/kg/min)** | 19.50 [16.30, 23.50] | 12.65 [11.12, 14.40] | <0.001 |
| **Peak $VO_2$ %** | 80.00 [68.73, 92.00] | 59.00 [49.25, 67.03] | <0.001 |
| **AT (ml/kg/min)** | 12.10 [10.50, 14.60] | 9.50 [8.40, 10.80] | <0.001 |
| **AT %** | 49.83 [42.04, 58.50] | 43.46 [34.05, 49.84] | <0.001 |
| **$VO_2$/work** | 8.98 ± 1.33 | 7.36 ± 1.64 | <0.001 |
| **Chronotropic index** | 47.07 [39.00, 61.00] | 59.65 [41.40, 88.63] | <0.001 |
| **Peak $O_2$ pulse (ml/beat)** | 11.30 [9.20, 14.20] | 7.90 [6.70, 9.60] | <0.001 |
| **Peak $O_2$ pulse %** | 89.00 [78.00, 103.00] | 71.00 [61.25, 84.50] | <0.001 |
| **$VE/VCO_2$ slope** | 33.50 [30.00, 38.00] | 46.00 [40.00, 53.75] | <0.001 |
| **$VEVCO_2/peakVO_2$** | 1.79 [1.27, 2.21] | 3.46 [2.97, 4.22] | <0.001 |
| **$P_{ET}CO_2$ (kPa)** | 4.44 ± 0.72 | 3.49 ± 0.65 | <0.001 |
| **Lactate (mmol/L)** | 7.71 ± 2.62 | 4.96 ± 1.83 | <0.001 |

| | | | |
|---|---|---|---|
| **A-a gradient (kPa)** | 3.95 [2.55, 5.98] | 6.31 [3.64, 8.24] | <0.001 |
| **$V_D/V_T$** | 0.30 ± 0.10 | 0.44 ± 0.09 | <0.001 |
| **NT-proBNP (pg/ml)** | 78.00 [40.50, 157.00] | 122.00 [62.00, 338.00] | 0.001 |
| **RVEF %** | 53.22 [44.97, 59.00] | 51.18 [42.26, 59.08] | 0.633 |

**Supplementary table 4. CPET parameters, MRI data and NT-proBNP based on $VEVCO_2/peakVO_2$ ≥ and < 2.7.**

*A-a gradient, alveolar-arterial oxygen difference; AT, anaerobic threshold; BR, breathing reserve; NT-proBNP, N-terminal brain natriuretic peptide; $O_2$, oxygen; $P_{ET}CO_2$, partial pressure of end-tidal carbon dioxide; RER, respiratory exchange ratio; RVEF, right ventricular ejection fraction; slope 1, slope of $O_2$ pulse with no warm-up phase; slope 2, slope of both warm-up and work phase; slope 3, slope of $O_2$ pulse in recovery; slope 4, slope of $O_2$ pulse before the plateau; $VE/VCO_2$, slope of ventilation to carbon dioxide production; $V_D/V_T$, physiological dead-space as a ratio to tidal volume; $VO_2$, oxygen uptake, $VO_2$/work, oxygen uptake per unit of work.*

| **Slope 1** | **< 0.40 (n=62)** | **≥ 0.40 (n=187)** | **p-value** |
|---|---|---|---|
| **AT (mg/kg/min)** | 10.57 ± 2.84 | 12.86 ± 4.27 | <0.001 |
| **AT (% predicted)** | 44.57 ± 12.33 | 51 ± 14.9 | 0.003 |
| **$VO_2$/work** | 7.36 ± 1.65 | 8.9 ± 1.45 | <0.001 |
| **Chronotropic Index** | 81.18 ± 27.07 | 48.03 ± 16.66 | <0.001 |
| **Peak $O_2$ pulse (ml/beat)** | 7.63 ± 2.29 | 11.78 ± 3.47 | <0.001 |
| **Peak $O_2$ pulse (% predicted)** | 68.42 ± 17.81 | 90.94 ± 20.93 | <0.001 |

| | | | |
|---|---|---|---|
| **$VEVCO_2/peakVO_2$** | 3.12 [2.35, 4.05] | 2.07 [1.4, 2.61] | <0.001 |
| **Peak $VO_2$ (ml/kg/min)** | 13.3 [11.5, 15.8] | 17.6 [14.6, 22.9] | <0.001 |
| **Peak $VO_2$ % predicted** | 59 [51, 69.7] | 76.1 [62.2, 90.5] | <0.001 |
| **$VE/VCO_2$ slope** | 42 [35, 51.2] | 35 [31, 41] | <0.001 |

**Supplementary table 5. CPET parameters based on slope 1 cut-off limit (≥ or < 0.40).**

*AT, anaerobic threshold; $O_2$, oxygen; Slope 1, slope of $O_2$ pulse with no warm-up phase; slope 2, slope of both warm-up and exercise; slope 3, slope of $O_2$ pulse in recovery; slope 4, slope of $O_2$ pulse before the plateau; $VE/VCO_2$, slope of ventilation to carbon dioxide production; $VO_2$, oxygen uptake, $VO_2$/work, oxygen uptake per unit of work.*

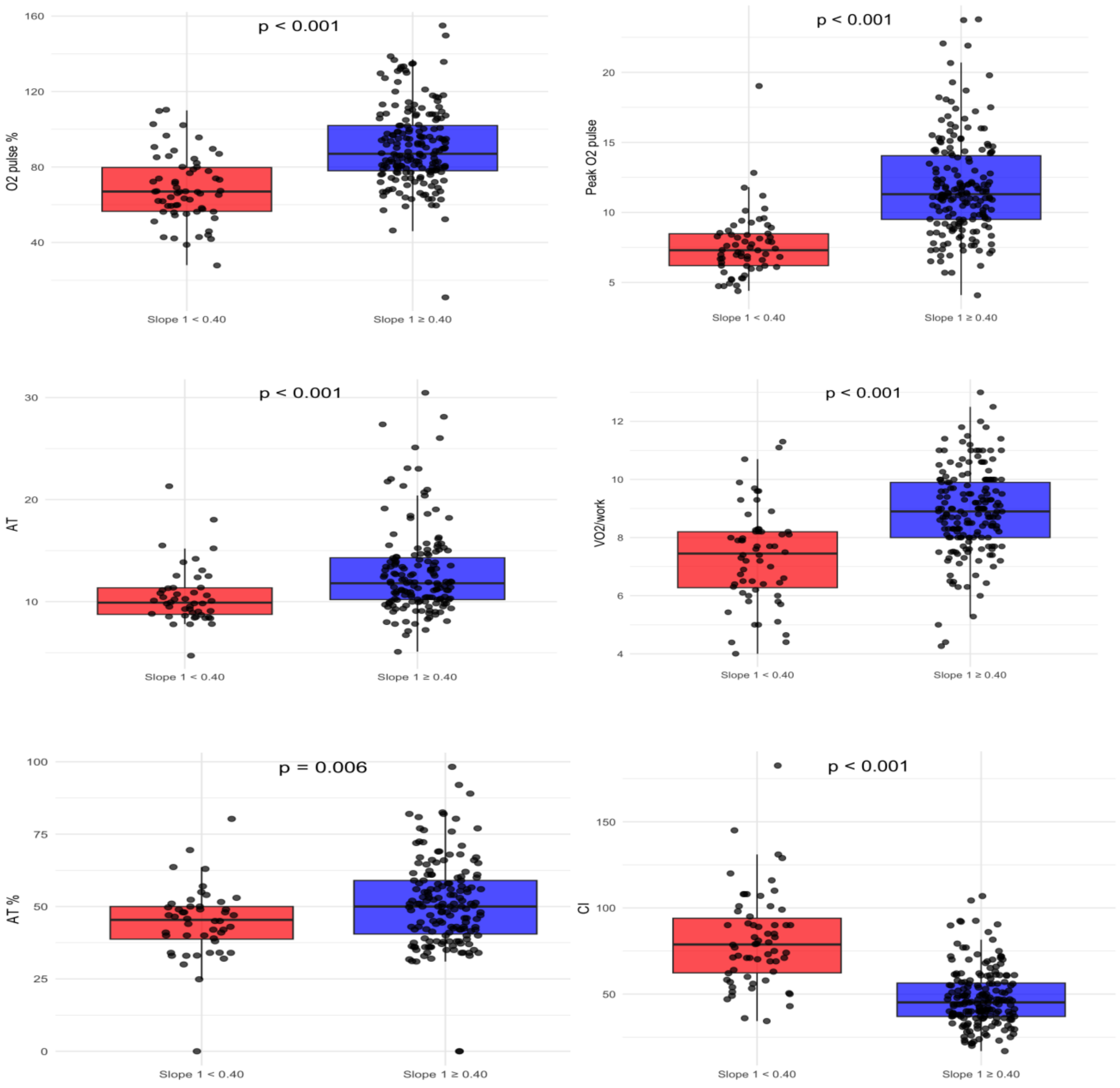


**Supplementary figure 4. Discrimination of oxygen delivery CPET parameters by slope 1.**

*AT, anaerobic threshold; AT %, anaerobic threshold as a percentage of maximum predicted oxygen uptake; CI, chronotropic index;* $O_2$*, oxygen; slope 1, slope of* $O_2$ *pulse with no warm-up phase;* $VO_2$*/work, oxygen uptake per unit of work*

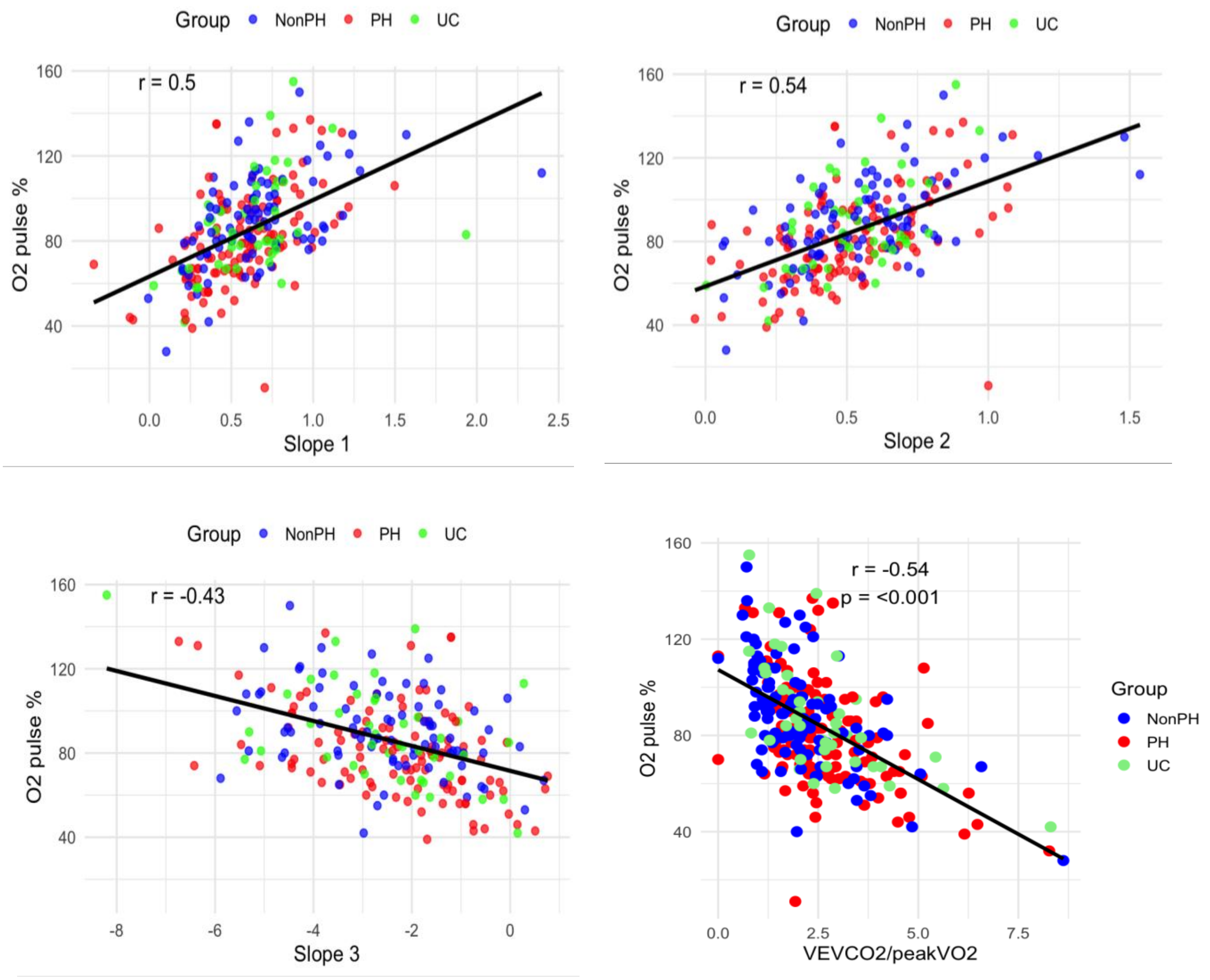


**Supplementary figure 5. Correlation of peak $O_2$ pulse % predicted with slopes and $VEVCO_2$/peak$VO_2$.**

*Slope 1, slope of $O_2$ pulse with no warm-up phase; slope 2, slope of both warm-up and work phase; slope 3, slope of $O_2$ pulse in recovery; VE/$VCO_2$, slope of ventilation to carbon dioxide production; $VO_2$, oxygen uptake.*

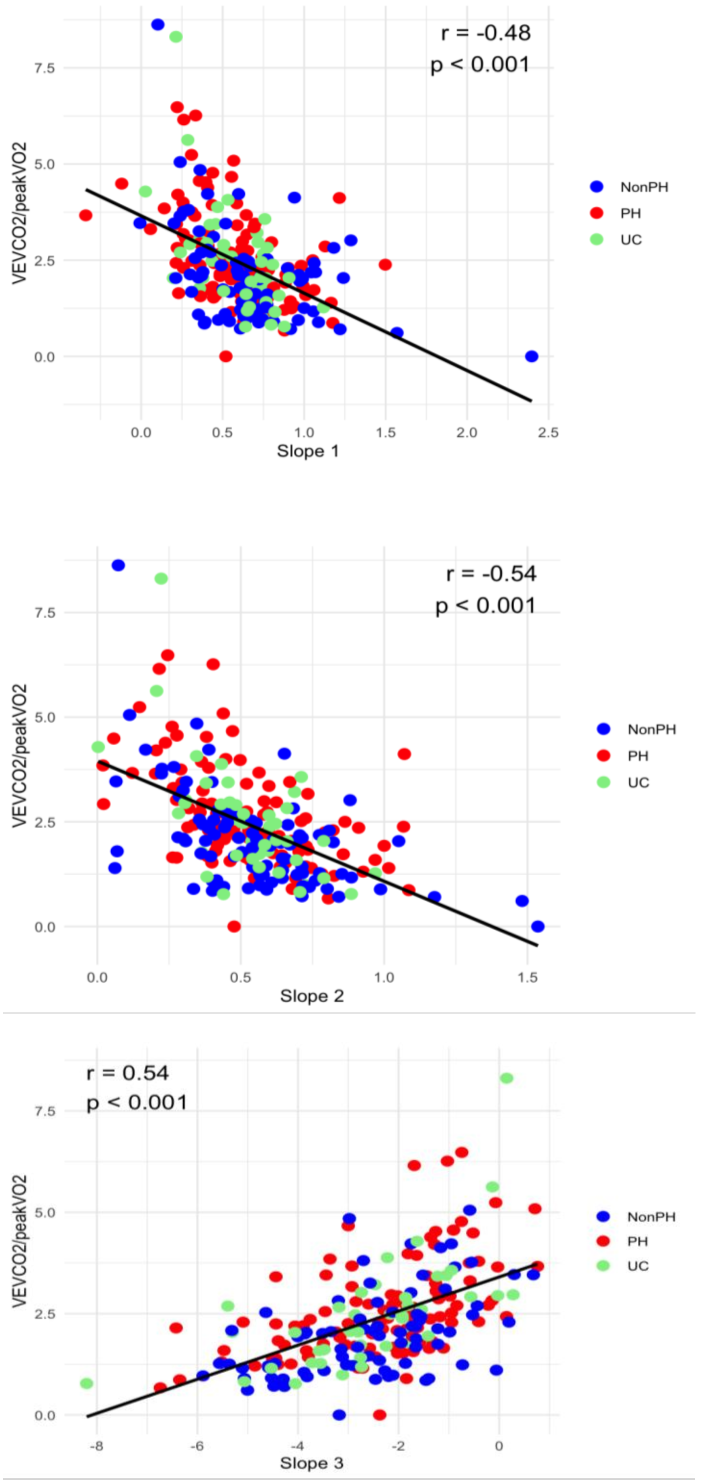


**Supplementary figure 6. Correlation of $VEVCO_2$/$peakVO_2$ with slopes.**

*Slope 1, slope of $O_2$ pulse with no warm-up phase; slope 2, slope of both warm-up and work phase; slope 3, slope of $O_2$ pulse in recovery.*

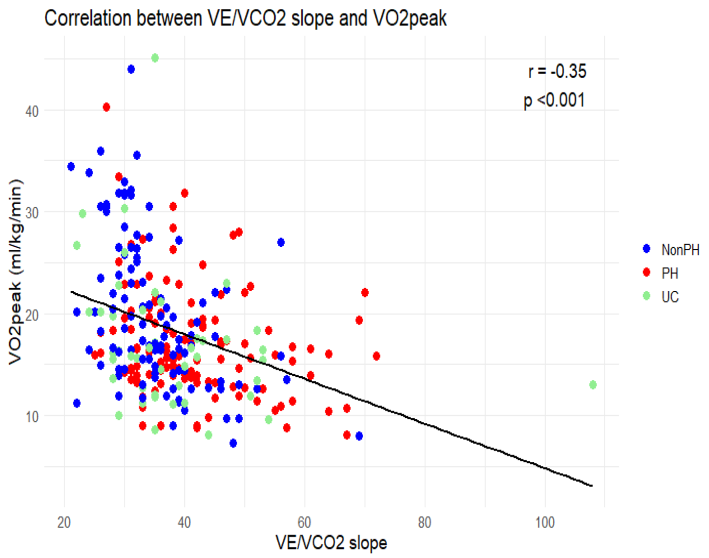


**Supplementary figure 7**. **Correlation between Peak $VO_2$ and $VE/VCO_2$ slope.**

*$VO_2$, oxygen uptake; $VE/VCO_2$, slope of ventilation to carbon dioxide production*.

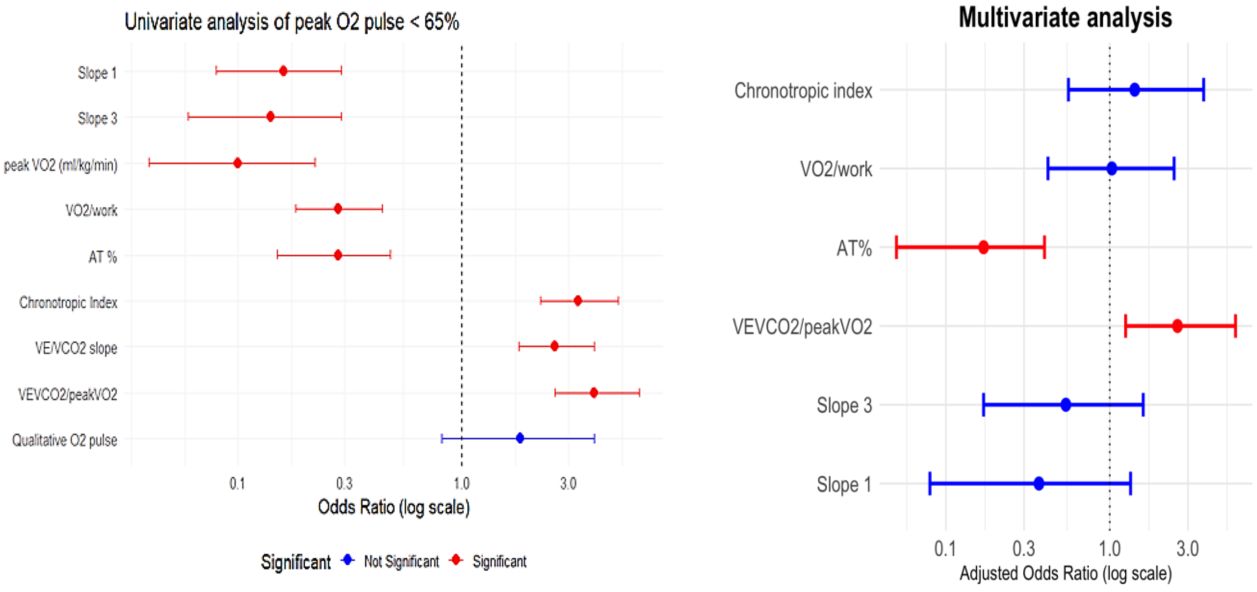


**Supplementary figure 8. Forest plots of univariable and multivariable analysis.**

*AT %, anaerobic threshold as a percentage of maximum predicted oxygen uptake; $O_2$, oxygen; slope 1, slope of $O_2$ pulse with no warm-up phase; slope 3, slope of $O_2$ pulse in recovery; $VE/VCO_2$, slope of ventilation to carbon dioxide production; $VO_2$, oxygen uptake; $VO_2$/work, oxygen uptake per unit of work.*

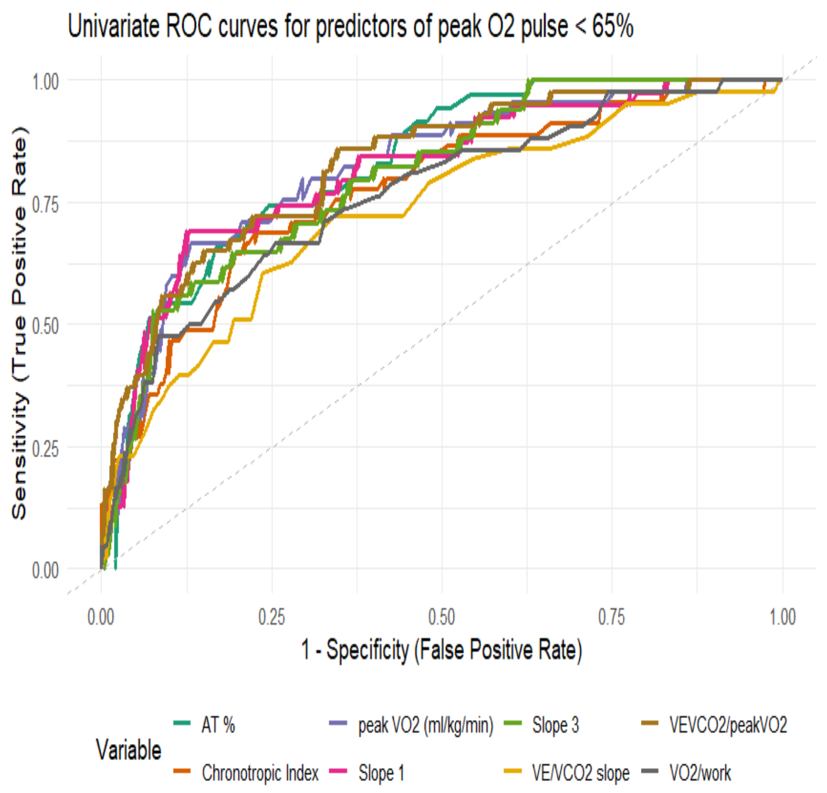


**Supplementary figure 9. ROC curves for oxygen delivery CPET parameters and slopes for the discrimination of peak $O_2$ pulse < 65% predicted.**

*AT %, anaerobic threshold as a percentage of maximum predicted oxygen uptake; slope 1, slope of $O_2$ pulse with no warm-up phase; slope 3, slope of $O_2$ pulse in recovery; VE/$VCO_2$, slope of ventilation to carbon dioxide production; $VO_2$, oxygen uptake; $VO_2$/work, oxygen uptake per unit of work.*

| Variable | OR (CI) | p-value |
|---|---|---|
| **Slope 1** | 0.37 (0.08-1.34) | 0.164 |
| **Slope 3** | 0.54 (0.17-1.60) | 0.278 |
| **$VEVCO_2$/peak$VO_2$** | 2.59 (1.25-5.85) | 0.014 |
| **AT%** | 0.17 (0.05-0.40) | $<0.001$ |
| **$VO_2$/work** | 1.03 (0.42-2.47) | 0.952 |
| **Chronotropic index** | 1.42 (0.56-3.74) | 0.463 |

**Supplementary table 6. Multivariable analysis.**

*AT, anaerobic threshold; slope 1, slope of $O_2$ pulse with no warm-up phase; slope 3, slope of $O_2$ pulse in recovery; VE/$VCO_2$, slope of ventilation to carbon dioxide production; $VO_2$, oxygen uptake, $VO_2$/work, oxygen uptake per unit of work.*

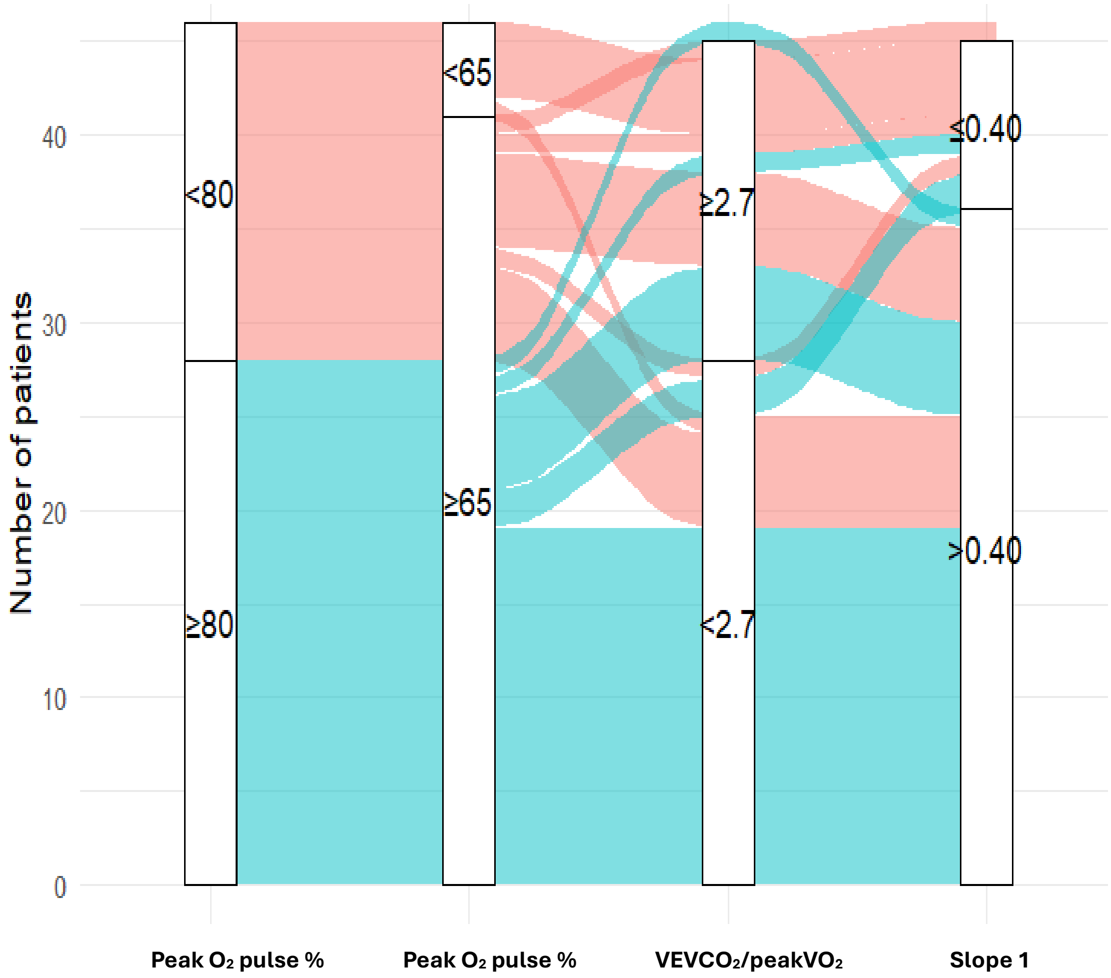


**Supplementary figure 10**. Sankey/alluvial graph showing the distribution of patients with unclassified pulmonary hypertension based on different markers of cardiac dysfunction.

*$O_2$, oxygen; slope 1, slope of $O_2$ pulse with no warm-up phase; VE/$VCO_2$, slope of ventilation to carbon dioxide production; $VO_2$, oxygen uptake. Pink= peak $O_2$ pulse <80%, green= peak $O_2$ pulse ≥ 80%.*
